# Comparison of different isolation periods for preventing the spread of COVID-19: a rapid systematic review and a modelling study

**DOI:** 10.1101/2023.01.12.23284479

**Authors:** Ya Gao, Yunli Zhao, Xi Zhang, Jinhui Tian, Gordon Guyatt, Qiukui Hao

## Abstract

**Background:** The optimal isolation duration for COVID-19 patients remains unclear. To support an update of WHO Living Clinical management guidelines for COVID-19 (https://www.who.int/publications/i/item/WHO-2019-nCoV-clinical-2022.2), this rapid systematic review and modelling study addresses the effects of different isolation periods for preventing onward transmission leading to hospitalization and death among secondary cases.

**Methods:** We searched World Health Organization (WHO) COVID-19 database for clinical studies evaluating the impact of isolation periods for COVID-19 patients up to July 28, 2022. We performed random-effects meta-analyses to summarize testing rates of persistent test positivity rates after COVID-19 infection. We developed a model to compare the effects of the five-day isolation and removal of isolation based on a negative antigen test with ten-day isolation on onward transmission leading to hospitalization and death. We assumed that patients with a positive test are infectious and those with a negative test are not. If the test becomes negative, patients will stay negative. The model included estimates of test positivity rates, effective reproduction number, and hospitalization rate or case fatality rate.

**Findings:** Twelve studies addressing persistent test positivity rates including 2799 patients proved eligible. Asymptomatic patients (27.1%, 95% CI: 15.8% to 40.0%) had a significantly lower rapid antigen test (RAT) positive rate than symptomatic patients (68.1%, 95% CI: 40.6% to 90.3%) on day 5. The RAT positive rate was 21.5% (95% CI: 0 to 64.1%; moderate certainty) on day 10. Our modelling study suggested that the risk difference (RD) for asymptomatic patients between five-day isolation and ten-day isolation in hospitalization (2 more hospitalizations of secondary cases per 1000 patients isolated, 95% uncertainty interval (UI) 2 more to 3 more) and mortality (1 more per 1000 patients, 95% UI 0 to 1 more) of secondary cases proved very small (very low certainty). For symptomatic patients, the potential impact of five- versus ten-day isolation was much greater in hospitalizations (RD 19 more per 1000 patients, 95% UI 14 more to 24 more; very low certainty) and mortality (RD 5 more per 1000 patients, 95% UI 4 more to 6 more; very low certainty). There may be no difference between removing isolation based on a negative antigen test and ten-day isolation in the onward transmission leading to hospitalization or death, but the average isolation period (mean difference −3 days) will be shorter for the removal of isolation based on a negative antigen test (moderate certainty).

**Interpretation:** Five versus 10 days of isolation in asymptomatic patients may result in a small amount of onward transmission and negligible hospitalization and mortality, but in symptomatic patients concerning transmission and resulting hospitalization and mortality. The evidence is, however, very uncertain.

**Funding:** WHO.

**Research in context:** *Evidence before this study:* Isolating infected patients and quarantining individuals with a high risk of recent infection remain widely used strategies to prevent the spread of SARS-CoV-2. There are no prior systematic reviews to evaluate effects relevant to decisions regarding protocols for ending COVID-19 isolation. Many modelling studies have, however, evaluated impact of five days of isolation or alternative strategies (e.g. 7 days and 10 days) with or without one negative lateral flow device on secondary infections or additional transmission risk. However, none has focused on the most patient-important outcomes - onward transmission leading to hospitalization or death. The optimal isolation duration for COVID-19 patients remains unclear. We searched WHO COVID-19 database for clinical studies evaluating the impact of isolation periods for COVID-19 patients up to July 28, 2022. We performed random-effects meta-analyses to summarize testing rates of persistent test positivity rates after COVID-19 infection. We used a model to compare the effects of the five-day isolation and removal of isolation based on a negative antigen test with ten-day isolation on onward transmission leading to hospitalization and death.

*Added value of this study:* To our knowledge, this is the first systematic review and modelling study to compare effects of the five-day isolation and removal of isolation based on a negative antigen test with ten-day isolation on most patient-important outcomes - onward transmission leading to hospitalization or death. This study demonstrates that for symptomatic patients the five-day isolation may increase onward transmission and thus hospitalization and mortality of secondary cases compared with the ten-day isolation by a magnitude most would consider important. For asymptomatic patients, the increase in hospitalizations and death may be small enough to be considered unimportant. Removal of isolation based on a negative antigen test will probably shorten the average isolation period compared with isolating all patients for 10 days.

*Implications of all the available evidence:* Our study provides evidence that 5 versus 10 days of isolation in asymptomatic patients may result in a small amount of onward transmission and negligible hospitalization and mortality, but in symptomatic patients concerning transmission and resulting hospitalization and mortality.

## Introduction

Coronavirus disease 2019 (COVID-19) has, as of October 23, 2022, resulted in over 624 million confirmed cases and more than 6.5 million deaths worldwide.^1^ Healthcare setting and community transmissions play an important role in the spread of the disease. Isolating infected patients and quarantining individuals with a high risk of recent infection remain widely used strategies to prevent the spread of severe acute respiratory syndrome coronavirus 2 (SARS-CoV-2).^2,3^

The current World Health Organization (WHO) recommendations, published on June 17 2020^4^ and September 15 2022^5^, for discharging patients from isolation differ for symptomatic and asymptomatic patients. For symptomatic patients, WHO recommends ten days after symptom onset plus at least three additional days without symptoms. For asymptomatic patients, WHO recommends ten days after a positive test for SARS-CoV-2.^4,5^

Since the COVID-19 outbreak, SARS-CoV-2 variants have changed from the initial outbreak strain to the delta variant and, most recently, the omicron variant. Compared to previous SARS-CoV-2 variants, the omicron variant is both more transmissible and has a shorter incubation period but has lower viral loads at diagnosis and a generally less severe course.^6-8^ Isolation or quarantine to limit its spread has high economic, societal, and psychological costs.^9-11^ These findings led the WHO to review its recommendations regarding isolation.

There are no prior systematic reviews to evaluate effects relevant to decisions regarding protocols for ending COVID-19 isolation. Many modelling studies have, however, evaluated impact of five days of isolation or alternative strategies (e.g. 7 days and 10 days) with or without one negative lateral flow device on secondary infections or additional transmission risk.^12-15^ However, none has focused on the most patient-important outcomes - onward transmission leading to hospitalization or death.

To support an update of WHO Living Clinical management guidelines for COVID-19 ^5^, we conducted a rapid systematic review and a modelling study. In the review, we evaluated SARS-CoV-2 testing positivity rates after isolation (i.e., 5 to 14 days) following diagnosis. In the modelling study, we evaluated the impact of five-day isolation, removal of isolation based on a negative antigen test, and ten-day isolation periods on onward transmission leading to hospitalization and death.

## Methods

This rapid systematic review adhered to the Cochrane guidance for rapid reviews^16^ and the Preferred Reported Items for Systematic Reviews and Meta-Analyses 2020 (PRISMA 2020) statement.^17^ We registered this rapid systematic review protocol with PROSPERO (CRD42022348626).

We conducted this review following the WHO predefined population, intervention, comparator, and outcome criteria: randomized controlled trials or observational studies that directly compared impact of five-day isolation and removal of isolation based on a negative antigen test with the current WHO-recommended isolation period of ten days for COVID-19 patients on onward transmission leading to hospitalization or death. Because we found no direct evidence addressing the question in either randomized trials or observational studies, we included evidence regarding SARS-CoV-2 testing positivity (i.e., viral culture, rapid antigen test, and PCR test) from 5 days after documented infection onward. On the basis of this evidence, and the best evidence regarding a model to estimate onward transmission leading to hospitalization or death.

### Eligibility criteria

We included clinical studies of any design with COVID-19 patients confirmed by PCR test or rapid antigen test addressing the impact of any isolation strategy on preventing the spread of COVID-19. There were no restrictions on publication language, publication status (peer-reviewed, in press, or preprint), age of patients, the severity of COVID-19, variants of SARS-COV-2, comorbidity of patients, isolation location, or co-interventions. We excluded studies enrolling people with suspected or probable COVID-19 (over 20% of participants) or contacts with confirmed COVID-19.

### Outcomes

The patient-important outcomes of interest were onward transmission leading to hospitalization or death. Viral culture positivity, rapid antigen test positivity, and PCR test positivity provided indirect evidence.

### Data sources and searches

With the aid of an expert librarian from the WHO, we searched the WHO COVID-19 database up to July 28, 2022. The WHO COVID-19 database is a comprehensive multilingual source of current literature on the topic, including global literature from over 25 bibliographic and grey literature sources. Appendix 1 presents the details of the search strategy.

### Study selection

We used Covidence (https://covidence.org/) for screening. Two reviewers independently screened titles and abstracts and subsequently the full texts of potentially eligible records. Reviewers resolved disagreements by discussion or, if necessary, by consultation with a third reviewer.

### Data extraction

Using a predesigned form, a reviewer conducted data extraction, and a second reviewer checked for the correctness and completeness of extracted data. Reviewers resolved discrepancies by discussion and, when necessary, with adjudication by a third reviewer. We extracted the following data: study characteristics (first author, study design, publication year, publication status, country, and sample size); patient characteristics (age, sex, severity of COVID-19, symptom status, vaccination status, and SARS-CoV-2 variant of concern); characteristics of isolation (isolation periods, isolation location, and co-interventions); and data on each outcome of interest.

### Risk of bias assessment

To assess the risk of bias in eligible studies regarding test positivity after infection, we used five domains of the Quality In Prognosis Studies (QUIPS) tool^18^: study participation, study attrition, prognostic factor measurement, outcome measurement, and statistical analysis and reporting. A reviewer rated each domain as either low, moderate, or high risk of bias. A second reviewer verified the judgments. Reviewers resolved discrepancies by discussion and, when necessary, with adjudication by a third reviewer. We considered studies were at overall low risk of bias if we judged four or more domains at low risk of bias; studies were at high risk of bias if we judged one or more domains at high risk of bias. We judged the remaining studies were at moderate risk of bias.

## Statistical analysis

### Systematic review analysis

The protocol included plans to pool hospitalization and mortality data (PROSPERO: CRD42022348626) but such direct evidence proved unavailable. Eligible studies reported the SARS-CoV-2 virus testing positivity rates following diagnosis. Using R (version 4.1.1, R Foundation for Statistical Computing) we performed meta-analyses to estimate proportions and associated 95% confidence intervals (CIs) using the restricted maximum likelihood (REML) method with the random-effects model.^19^ We used Freeman-Tukey double arcsine transformation to stabilize variances.^20^ We assessed the between-study heterogeneity with a visual inspection of forest plots and the I^2^ statistic.

### Modelling study

To estimate the impact of different isolation strategies on patient import outcomes (i.e., onward transmission leading to hospitalization and death), we first performed a modelling study and further conducted a more complex microsimulation modelling study as a sensitivity analysis. We modelled three different strategies to end COVID-19 isolation: five-day isolation (that is, patients with COVID-19 are isolated for five days, then can end isolation without any further consideration); removal of isolation based on negative antigen test (that is, patients with COVID-19 are isolated and receive a rapid antigen test daily from day five to day nine, those who test negative can end isolation while those who test positive continue to isolate until test negative or day ten); ten-day isolation (that is, patients with COVID-19 are isolated for ten days, then can end isolation without any further consideration). We compared the impact of the five-day isolation and removal of isolation based on negative antigen test with ten-day isolation on onward transmission leading to hospitalization and death.

We modelled a sample of 1000 individuals with confirmed COVID-19. To estimate the hospitalization and death for secondary cases of five-day isolation and ten-day isolation, we used an effective secondary reproduction number of 0.96 (95% CI 0.72 to 1.2),^21^ a hospitalization rate of 4.3%,^22^ and a case fatality rate of 1.05%^1^. To calculate 95% uncertainty intervals (UIs), we used the 95% CIs of the effective secondary reproduction number. We assumed that patients with a positive test (rapid antigen, viral culture) are infectious and those with a negative test are not. Further, if the test becomes negative, patients will stay negative. We used multiplication equations (test positivity per 1000 patients multiplied by effective reproduction number and hospitalization rate or case fatality rate) to estimate the number of hospitalization and death for secondary cases.

Given the assumption that patients with a negative rapid antigen test are non-infectious, onward transmission, hospitalization, and death using strategies of terminating isolation at the first negative antigen test will be identical to the ten-day isolation strategy. To calculate the average isolation period associated with the strategy of isolation terminated with the first negative test, we used the following equation:

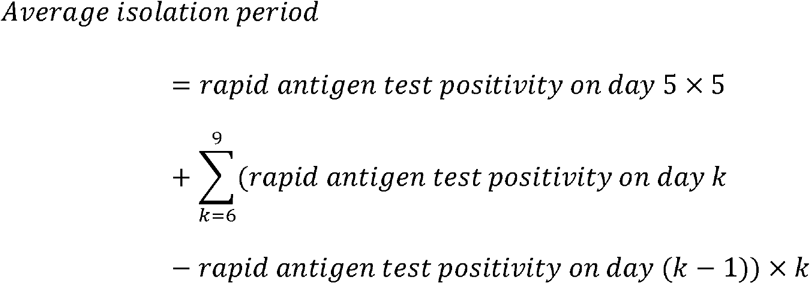

As a sensitivity analysis, we performed a more complex microsimulation modelling study using the model developed by Quilty and colleagues^23^, revised to suit our study purposes. In the complex microsimulation model, using a baseline Ct level of 40, incubation period of 3.42 days,^24^ a peak Ct value of 22.3 for symptomatic individuals,^25^ viral shedding time for symptomatic infections of 19.7 days (95%CI 17.2 to 22.7), asymptomatic infections of 10.9 days (95% CI 8.3 to 14.3 days),^26^ and day 5, day 6, and day 10 rapid antigen test positivity or viral culture positivity data from our rapid systematic review, we simulated a viral load trajectory of Ct values over the course of infection for each patient. We assumed that if the Ct value is less than 30, the individual is infectious.^27^ Appendix 2 presents the detailed methods.

### Subgroup analysis

As requested by the WHO guideline panel, we performed prior-specified subgroup analyses by symptom status (asymptomatic versus symptomatic patients with a prior subgroup hypothesis that asymptomatic patients would have a lower positive rate) and vaccination status, though evidence of rapid antigen test was unavailable for the latter analysis. If at least two studies provided information on a subgroup, we performed within-study subgroup analyses. To assess the credibility of significant subgroup effects, we used a version of the Instrument for assessing the Credibility of Effect Modification Analyses (ICEMAN) tool, originally developed for randomized trials and meta-analysis of randomized trials, modified for the issue of test positivity over the post-infection period.^28^ A finding of moderate or highly credible subgroup effects mandated a focus on subgroup results to inform the WHO panel’s recommendations.

### Certainty of evidence

For results of meta-analyses of SARS-CoV-2 testing positivity rates following diagnosis, we used the GRADE (Grading of Recommendations Assessment, Development, and Evaluation) approach for overall prognosis in broad populations^29^ to rate the overall certainty of the evidence for each outcome as ‘‘very low’’, ‘‘low’’, ‘‘moderate’’, or ‘‘high’’. The assessment included five domains: risk of bias^30^, imprecision^31^, inconsistency^32^, indirectness^33^, and publication bias^34^. To assess the certainty of the evidence from our modelling study, we used criteria adapted from GRADE guidelines for assessing the certainty of modelled evidence.^35^

## Results

Systematic review of clinical studies

### Study identification

The electronic database search identified 2408 records. After screening 2228 titles and abstracts and 30 full texts, 12 studies^36-47^ proved eligible (Figure 1).

**Figure 1.**
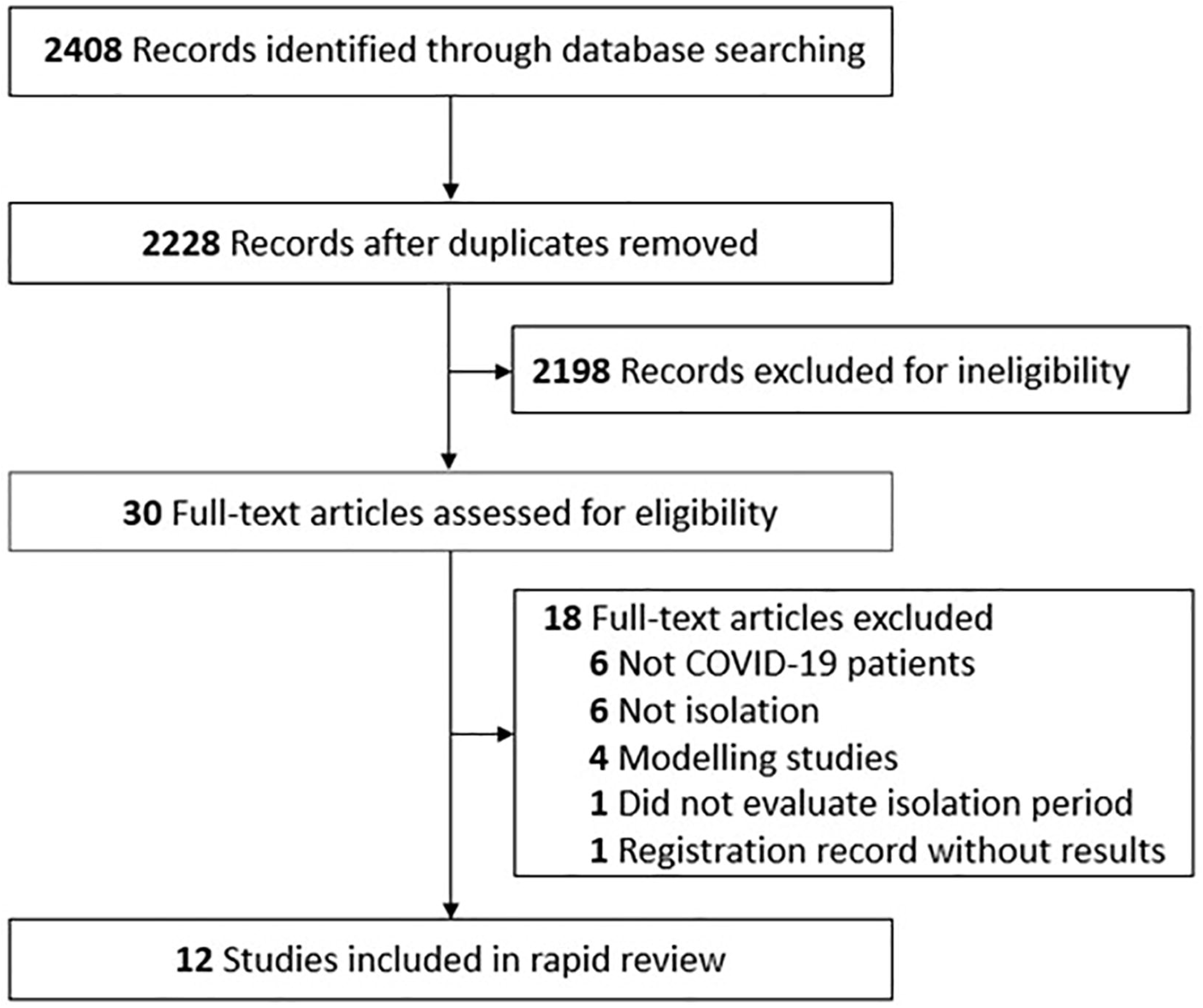
Flow diagram of study selection.

### Characteristics of eligible studies

Table 1 summarizes the 12 cohort studies that proved eligible, all of which were published in 2022 and addressed COVID-19 positivity from 5 to 14 days after diagnosis. The sample size ranged from 10 to 729 (a total of 2799) and the proportion of males from 36.7% to 90.0%. Most studies included patients with omicron SARS-CoV-2 variants, although seven studies did not provide the distribution of omicron variants. Two studies included only asymptomatic patients, two only symptomatic patients, six both asymptomatic and symptomatic patients, and two did not provide relevant information.

**Table 1.**
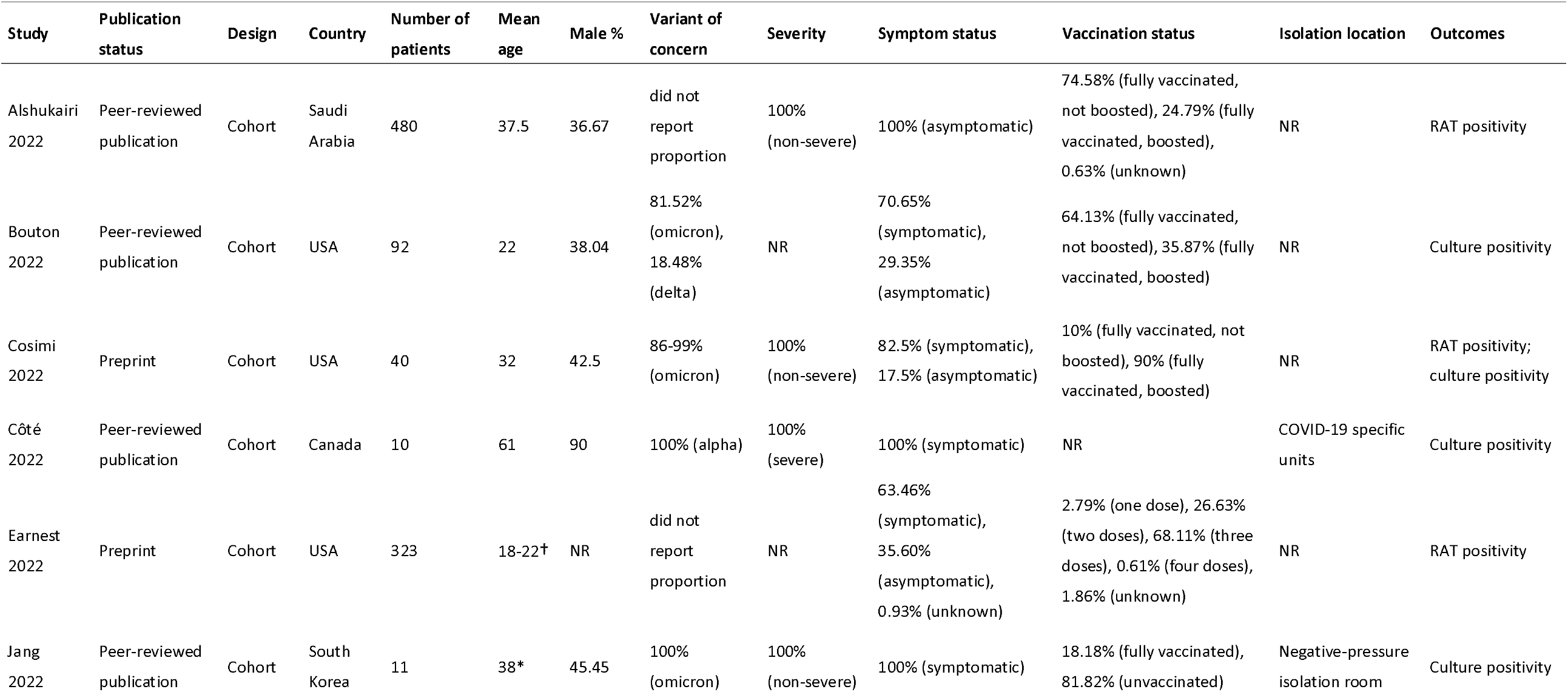
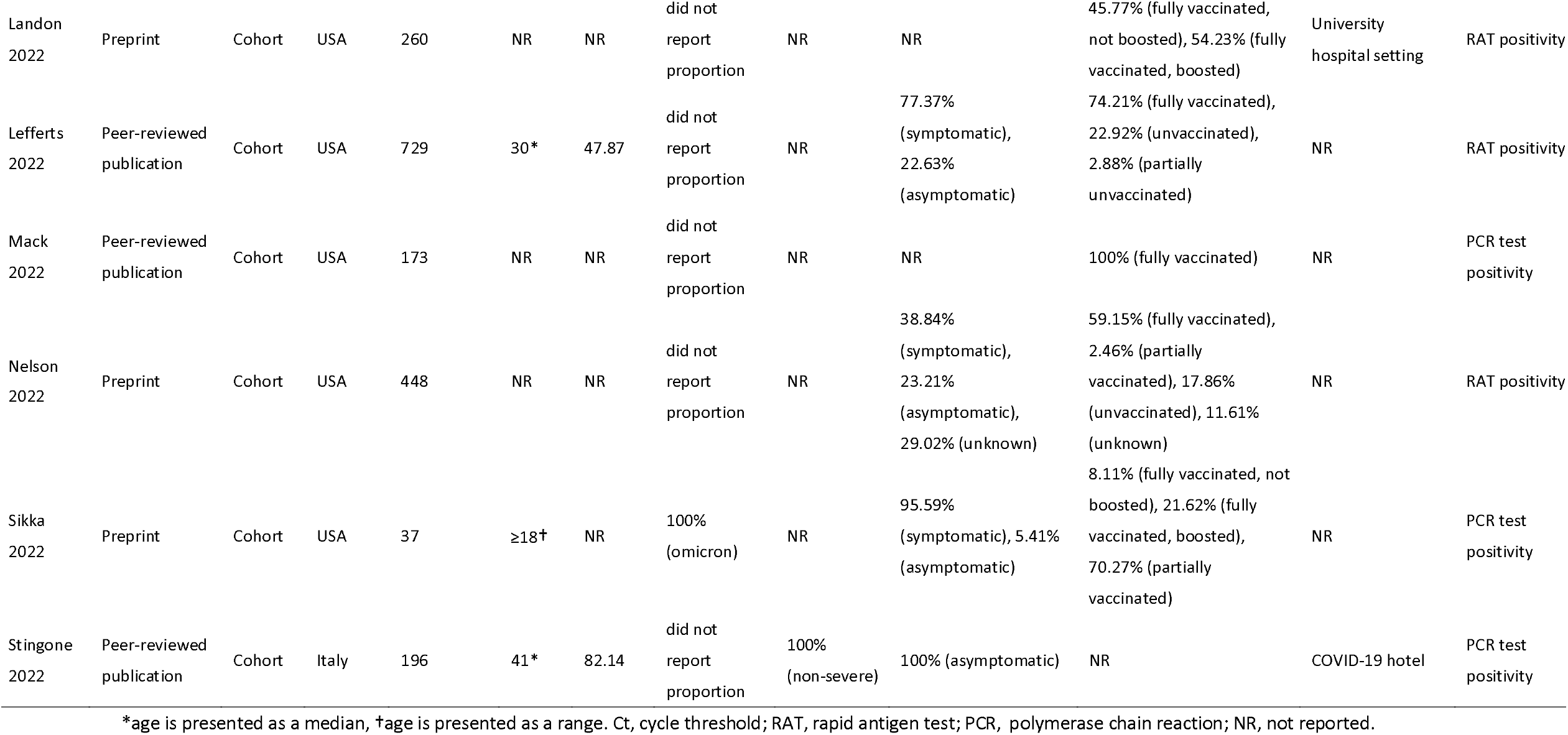
Characteristics of eligible studies.

### Risk of bias

Eight studies^36-38,40,43,45-47^ had low risk of bias and four^39,41,42,44^ had moderate risk of bias. Most biases were due to the lack of clear definition or description of the prognostic factor (symptom status) and poor reporting of the statistical analysis (Appendix 3).

### Outcomes

Moderate certainty evidence showed the pooled percentage of patients with positive rapid antigen test was 48.3% (95%CI 34.2% to 62.5%; 4 studies^40,42,43,45^, 667 patients) on isolation day 5; 47.5% (95%CI 28.2% to 67.1%; 5 studies^38,40,42,43,45^, 691 patients) on day 6; and 21.5% (95%CI 0% to 64.1%; 3 studies^38,40,42^, 368 patients) on day 10 (Appendix 4 and Appendix 5). Within-study subgroup analyses (Figure 2 and Appendix 6) showed significant subgroup effects between asymptomatic patients and symptomatic patients on day 5 (pooled ratio of percentage is 0.39, 95% CI: 0.27 to 0.57, P for interaction < 0.001) and day 6 (pooled ratio of percentage is 0.47, 95% CI: 0.32 to 0.68, P for interaction < 0.001) rapid antigen test positivity. We judged the credibility of these subgroup effects as moderate (Table 2). Asymptomatic patients had a lower rapid antigen test positive rate than symptomatic patients from day 5 to day 9 (Figure 3).

**Table 2.**
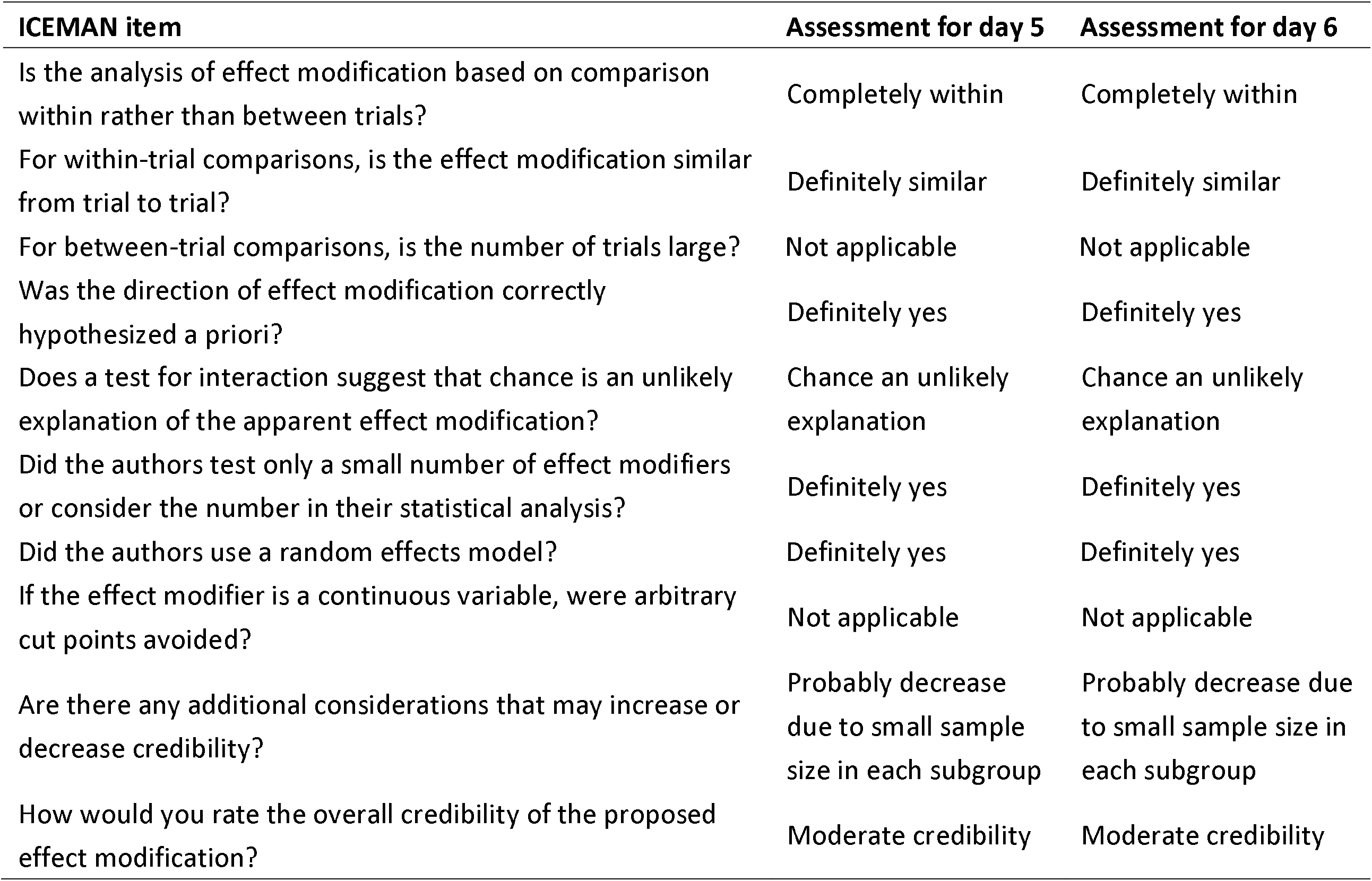
Credibility assessment of subgroup analysis for day 5 and 6 rapid antigen test positivity by symptom status.

**test positivity by symptom status**
| **ICEMAN item** | **Assessment for day 5** | **Assessment for day 6** |
| --- | --- | --- |
| Is the analysis of effect modification based on comparison within rather than between trials? | Completely within | Completely within |
| For within-trial comparisons, is the effect modification similar from trial to trial? | Definitely similar | Definitely similar |
| For between-trial comparisons, is the number of trials large? | Not applicable | Not applicable |
| Was the direction of effect modification correctly hypothesized a priori? | Definitely yes | Definitely yes |
| Does a test for interaction suggest that chance is an unlikely explanation of the apparent effect modification? | Chance an unlikely explanation | Chance an unlikely explanation |
| Did the authors test only a small number of effect modifiers or consider the number in their statistical analysis? | Definitely yes | Definitely yes |
| Did the authors use a random effects model? | Definitely yes | Definitely yes |
| If the effect modifier is a continuous variable, were arbitrary cut points avoided? | Not applicable | Not applicable |
| Are there any additional considerations that may increase or decrease credibility? | Probably decrease due to small sample size in each subgroup | Probably decrease due to small sample size in each subgroup |
| How would you rate the overall credibility of the proposed effect modification? | Moderate credibility | Moderate credibility |

**Figure 2.**
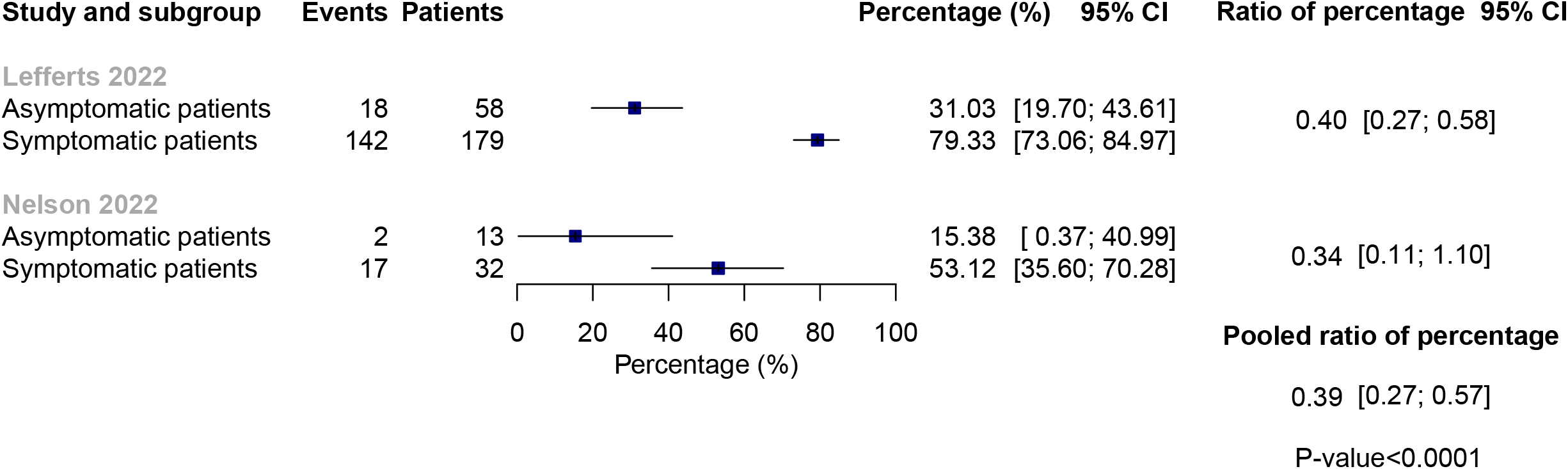
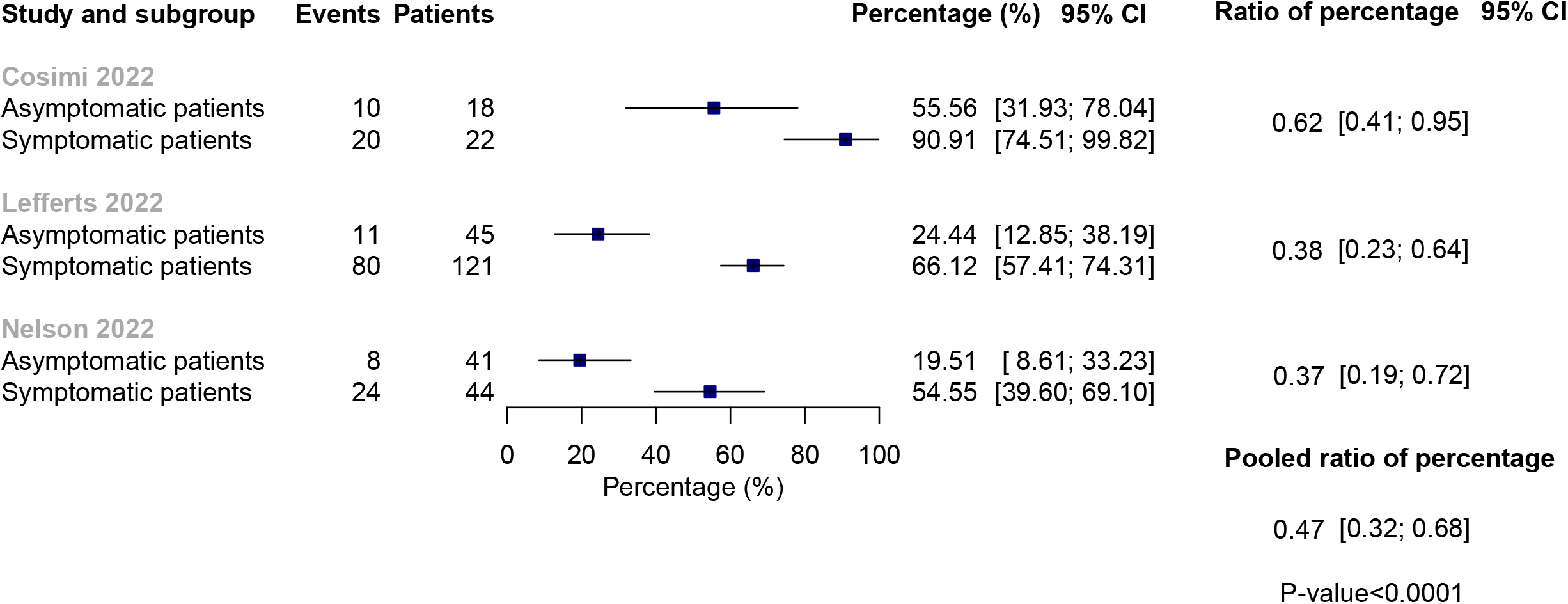
Within-study subgroup analysis of day 5 and day 6 rapid antigen test positivity by symptom status. (A) day 5; (B) day 6.

**Figure 3.**
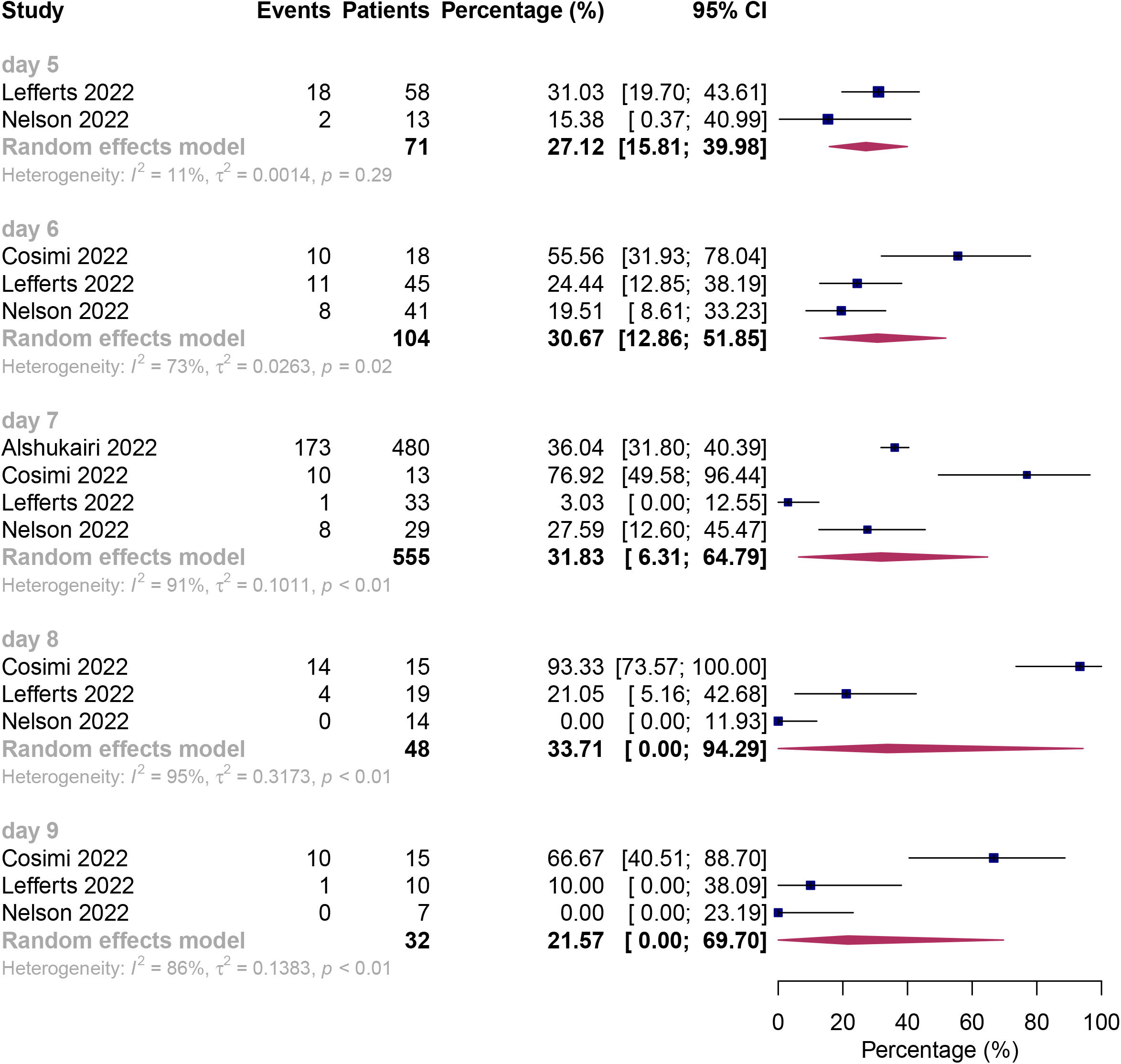
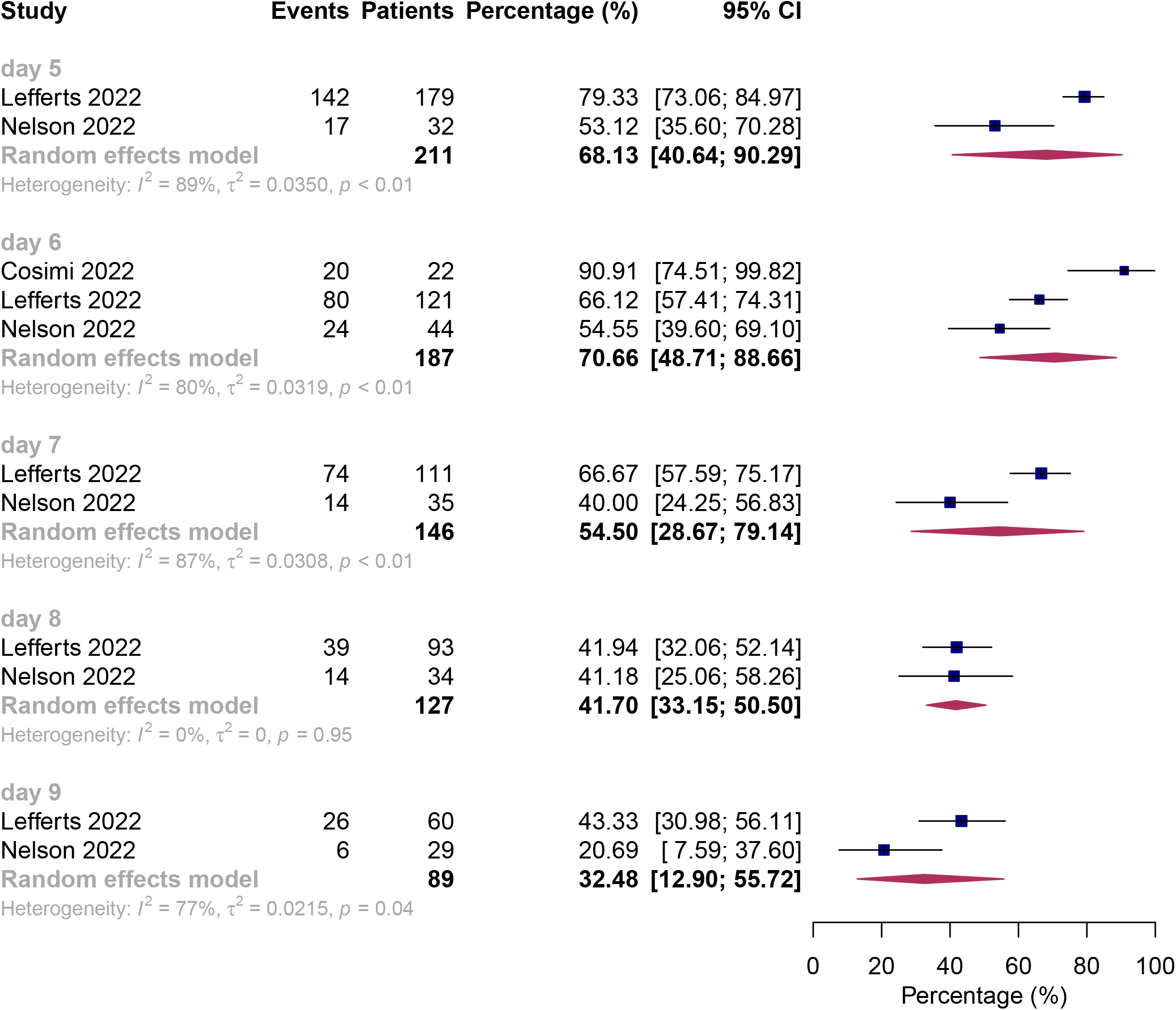
Pooled percentage of rapid antigen test positivity for asymptomatic and symptomatic patients. (A) asymptomatic patients; (B) symptomatic patients.

Low certainty evidence revealed that the percentage of patients with positive viral cultures was 63.6% (95%CI 32.7% to 90.0%; 1 study^41^, 11 patients) on day 5; 35.3% (95%CI 5.0% to 73.5%; 3 studies^37,38,41^, 120 patients) on day 6; and 0% (95%CI 0% to 15.1%; 1 study^41^, 11 patients) on day 10 (Appendix 5 and Appendix 7). Between-study subgroup analyses indicated that, on day 6, asymptomatic patients had a lower culture-positive rate (22.2% versus 63.5%, P for interaction = 0.05) than symptomatic patients, and fully vaccinated patients had a significantly lower culture-positive rate (13.7% versus 88.9%, P for interaction < 0.01) than unvaccinated patients (Appendix 8). However, due to the between-study comparison, small sample size, and chance remaining a likely explanation for findings, we rated the credibility of these subgroup effects as low (Appendix 9). Appendix 10 presents additional results that include PCR test positivity data.

### Modelling studies

Our model results suggested, for asymptomatic patients, there may be a small difference between five-day isolation and ten-day isolation in 28-day hospitalization (risk difference (RD) 2 more hospitalizations of secondary cases per 1000 patients isolated, 95% UI 2 more to 3 more) and 90-day mortality (RD 1 more death of secondary cases per 1000 patients isolated, 95% UI 0 to 1 more). For symptomatic patients, the five-day isolation may increase hospitalization (RD 19 more hospitalizations of secondary cases per 1000 patients isolated, 95% UI 14 more to 24 more) and mortality (RD 5 more deaths of secondary cases per 1000 patients isolated, 95% UI 4 more to 6 more) compared with the ten-day isolation (Table 3). Appendix 11 presents the GRADE summary of findings for outcomes estimated from positive viral culture data.

**Table 3.** GRADE summary of findings.

| Outcome | Absolute effect estimates |  | Certainty of the evidence | Plain language summary |
| --- | --- | --- | --- | --- |
| 3.1 Isolation for 5 days versus isolation for 10 days |  |  |  |  |
|  | Isolation for 5 days | Isolation for 10 days |  |  |
| 3.1.1 All patients |  |  |  |  |
| Onward transmission leading to hospitalization (28 days) | 20<br>per 1000 | 9<br>per 1000 | Very low<br>Due to certainty of parameters (moderate) in the model and indirectness | Whether isolation of 5 days compared with 10 days would increase onward transmission leading to hospitalization for secondary cases is very uncertain. |
|  | Difference: 11 more per 1000<br>(95% UI 8 more to 14 more) |  |  |  |
| Onward transmission leading to death (90 days) | 5<br>per 1000 | 2<br>per 1000 | Very low<br>Due to certainty of parameters (moderate) in the model and indirectness | Whether isolation of 5 days compared with 10 days would increase onward transmission leading to death for secondary cases is very uncertain. |
|  | Difference: 3 more per 1000<br>(95% UI 2 more to 3 more) |  |  |  |
| 3.1.2 Symptomatic patients |  |  |  |  |
| Onward transmission leading to hospitalization (28 days) | 28<br>per 1000 | 9<br>per 1000 | Very Low<br>Due to certainty of parameters (moderate) in the model and indirectness | Whether isolation for 5 days compared with 10 days would increase onward transmission leading to hospitalization of secondary cases is very uncertain. |
|  | Difference: 19 more per 1000<br>(95% UI 14 more to 24 more) |  |  |  |
| Onward transmission leading to death (90 days) | 7<br>per 1000 | 2<br>per 1000 | Very Low<br>Due to certainty of parameters (moderate) in the model and indirectness | Whether isolation for 5 days compared with 10 days would increase onward transmission leading to death of secondary cases is very uncertain. |
|  | Difference: 5 more per 1000<br>(95% UI 4 more to 6 more) |  |  |  |
| 3.1.3 Asymptomatic patients |  |  |  |  |
| Onward transmission leading to hospitalization (28 days) | 11<br>per 1000 | 9<br>per 1000 | Very Low<br>Due to certainty of parameters (moderate) in the model and indirectness | Whether isolation for 5 days compared with 10 days would increase onward transmission leading to hospitalization of secondary cases is very uncertain. |
|  | Difference: 2 more per 1000<br>(95% UI 2 more to 3 more) |  |  |  |
| Onward transmission leading to death (90 days) | 3<br>per 1000 | 2<br>per 1000 | Very Low<br>Due to certainty of | Whether isolation for 5 days compared with 10 days would |
|  | Difference: **1 more per 1000**<br>(95% UI 0 to 1 more) |  | parameters (moderate) in the model and indirectness | increase onward transmission leading to death of secondary cases is very uncertain. |
| 3.2 Removal of isolation based on a negative antigen test versus isolation for 10 days |  |  |  |  |
|  | Removal of isolation based on a negative antigen test | Isolation for 10 days |  |  |
| Onward transmission leading to hospitalization (28 days) | 9 per 1000 | 9 per 1000 | Very Low<br>Due to certainty of parameters (moderate) in the model and indirectness | Whether removing isolation based on a negative antigen test compared with isolation of 10 days would increase onward transmission leading to hospitalization of secondary cases is very uncertain. |
|  | Identical |  |  |  |
| Onward transmission leading to death (90 days) | 2 per 1000 | 2 per 1000 | Very Low<br>Due to certainty of parameters (moderate) in the model and indirectness | Whether removing isolation based on a negative antigen test compared with isolation of 10 days would increase onward transmission leading to death of secondary cases is very uncertain. |
|  | Identical |  |  |  |
| Average isolation period | 7 days | 10 days | Moderate<br>Due to parameters in the model | Removing isolation based on the negative antigen test probably decreases average isolation compared with isolation for 10 days. |
|  | Mean difference: 3 days lower |  |  |  |

UI, uncertainty interval. Model assumptions: We assume that all patients with a negative rapid antigen test are non-infectious and assume if the test is negative, the patients would stay negative status.

There may be no difference in the hospitalization and mortality for secondary cases between removing isolation based on a negative antigen test before ten days and at the ten-day isolation (very low certainty), but the average isolation period (mean difference −3 days) will probably be shorter for the removal of isolation based on a negative antigen test compared with ten-day isolation (moderate certainty, Table 3). Most estimates from the modelling study are very low certainty.

Our sensitivity analysis using a more complex microsimulation model provided similar results in onward transmission leading to hospitalization (RD varied from 12 more to 17 more per 1000 patients isolated) and death (RD varied from 5 more to 7 more per 1000 patients isolated) of five-day isolation versus ten-day isolation for overall patients (Appendix 12.1). In the complex microsimulation model using assumptions different from our relatively simple model, whether removal of isolation based on a negative antigen test would increase hospitalization and mortality for secondary cases is very uncertain compared with isolation of 10 days (Appendix 12.2).

## Discussion

In this rapid systematic review of 12 cohort studies including 2799 COVID-19 patients, we found that the pooled percentage of rapid antigen test positivity on day 5 was 48.3% and asymptomatic patients had a lower rapid antigen test positive rate than symptomatic patients (27.1% versus 68.1%) that chance could not easily explain and that met ICEMAN criteria for a moderate credibility subgroup analysis. The percentage of positive cases decreased over the isolation time reaching, for the entire group, 21.5% on day 10.

Our primary modelling study yields very low certainty of evidence on onward transmission leading to hospitalization or death. It does suggest, however, that for symptomatic patients the five-day isolation may increase onward transmission and thus hospitalization and mortality of secondary cases compared with the ten-day isolation by a magnitude most would consider important (Table 3). For asymptomatic patients, the increase in hospitalizations and death may be small enough to be considered unimportant. A second modelling study based on alternative methods showed similar results. Removal of isolation based on a negative antigen test will probably shorten the average isolation period compared with isolating all patients for 10 days.

### Strengths and limitations

Our review is limited in that no studies, either randomized or observational, have directly addressed the impact of variable durations of isolation on forward transmission and its important possible consequences on hospitalization and mortality. We therefore had to address the issue with indirect evidence that informed our modelling study. This modelling study has substantial uncertainties with respect to several parameters including effective reproduction number, hospitalization rate, and case fatality rate. Some reassurance regarding the credibility of the results comes, however, from similar results in an alternative modelling approach.

In contrast to previous attempts to model the possible consequences of isolation periods, we conducted a systematic review of one key aspect of the relevant indirect evidence: the SARS-CoV-2 virus test positivity from 5 to 14 days after diagnosis. The strengths of this review include a comprehensive search, duplicate assessment of eligibility, independent checking of data abstraction, and appropriate statistical analysis, including the approach to addressing a key subgroup hypothesis: results confirmed the hypothesized markedly greater test positivity rates at five days in those with or without symptoms.

Forty-two percent of included studies were preprints that have not been peer-reviewed, and results may differ from the final published version, although the likelihood of results changing is low.^48^. There was substantial heterogeneity across studies for most meta-analyses. The severity of COVID-19, age, co-intervention, and immunosuppression status may be the critical factors attributed to the differences. However, the data to address subgroup analyses for these factors proved unavailable. Because the data were very limited, we could not perform meta-analyses regarding PCR test positivity rate.

To provide evidence of patient-important outcomes, we further performed modelling studies relying on the results of our review of test positivity studies. Strengths of our model include its transparency. To reflect the currently dominant variant of concern, for input parameters, we used data specific to the Omicron variant when available. Finally, our relatively simple model provided results similar to applying our parameter estimates to a more complex microsimulation model.

The main limitation of the model is uncertainty around key parameters, in particular reproduction number and hospitalization rate. These include some parameters (e.g., peak Ct value, duration of viral shedding) in which data specific to the Omicron variant were not available, requiring use of data from the overall SARS-COV-2 variants. Owing to limited data, we were unable to estimate outcomes for subgroups of patients (e.g., vaccination status). These limitations led us to classify the evidence regarding hospitalization and death as very low certainty.

### Comparisons with other studies

A randomised, controlled, non-inferiority trial reported that daily lateral flow device testing with 24h exemption from self-isolation appeared to be non-inferior to standard self-isolation (10 days) in reducing onward transmission of SARS-CoV-2 for contacts of confirmed COVID-19 cases.^49^ Another cluster-randomised, controlled trial reported that daily contact testing of school-based contacts was non-inferior to self-isolation of 10 days for control of COVID-19 transmission.^50^ These findings are consistent with our results on the comparison of removal of isolation based on negative antigen testing and ten-day isolation in confirmed COVID-19 patients. Implications for practice and future research Healthcare public policy decisions must rely on the best evidence, even if that evidence is very low quality. Recommendations regarding isolation after COVID-19 diagnosis, because of their major implications for large numbers of people, represent a compelling example. Despite limitations of the evidence regarding the magnitude of hospitalization resulting from 5 versus 10 days isolation, because of the moderate credibility of our analysis regarding duration of test positivity related to symptomatic status, there is a high likelihood that shorter isolation will result in appreciably less hospitalization in the asymptomatic than the symptomatic. The WHO panel using the results of our work was therefore able to recommend 5 days of isolation for asymptomatic and 10 days for symptomatic patients.

Our studies identify major gaps in the evidence regarding an optimal isolation period. Compelling evidence regarding optimal isolation will require conduct of randomized controlled trials. Future clinical studies could focus on patient-important outcomes (e.g. onward transmission leading to hospitalization and/or death) and test important subgroup hypotheses such as the variant of virus, disease severity, symptom status, vaccination status, immunosuppression status, and co-interventions.

## Conclusions

Our findings show that a five-day isolation may result in minimal hospitalization or death as a result of spread of COVID-19 for asymptomatic patients; the same may not be true for symptomatic patients. Removing isolation based on a negative antigen test is likely to shorten the average isolation period, possibly without negative consequences on onward transmission. Providing higher certainty evidence on the impact of alternative isolation strategies will require future studies using robust designs.

## Data Availability

Data in this systematic review with meta-analysis are extracted from published studies available on the internet. All processed data are presented in this article and the appendix.

## Contributors

YG, GG, and QH conceived and designed the study. YG and YZ screened and selected the articles. YG and YZ extracted the data and assessed the risk of bias. YG, XZ, and QH led the development and analysis of the model. GG supervised the data analyses. YG and QH rated the certainty of evidence. GG provided methodological support. YG, YZ, XZ, GG, JT, and QH interpreted the data. YG and QH drafted the manuscript. All authors reviewed the manuscript and approved the final version of the manuscript. YG and QH accessed and verified the underlying data. All authors had full access to all the data in the study and had final responsibility for the decision to submit for publication.

## Patient consent

Not required.

## Ethical approval

Not required.

## Acknowledgements

We thank Kavita Kothari (Health Information Specialist, WHO Library and Digital Information Networks, Geneva, Switzerland) for helping to conduct the searches. We thank Ahmed M Bayoumi (Institute of Health Policy, Management and Evaluation, University of Toronto, Toronto, ON, Canada) for contributing to the development of model. We thank Janet Diaz (World Health Organization, Geneva, Switzerland), John Adabie Appiah (Komfo Anokye Teaching Hospital, Kwame Nkrumah University of Science and Technology, Kumasi, Ghana), Srinivas Murthy (Division of Pediatrics, University of British Columbia, Vancouver, BC, Canada), and Julie Viry (World Health Organization, Geneva, Switzerland) for internal review of the manuscript. YG and YZ acknowledge funding from China Scholarship Council.

