## Appendix for "Comparison of different isolation periods for preventing the spread of COVID-19: a rapid systematic review and a modelling study"

### Appendix 1: Search strategy of WHO COVID-19 database

| **#** | **Search** |
| --- | --- |
| 1 | ("infective duration"~5 OR "contagious duration"~5 OR "contagiousness duration" OR "infectious duration"~5 OR "infectiousness duration"~5 OR "viral duration"~3 OR "CT value" OR "CT values" OR "cycle threshold value" OR "cycle threshold values" OR "spread duration"~5 OR "infectivity duration"~5 OR "infective period"~5 OR "contagious period"~5 OR "infectious period"~5 OR "CT value*" OR "cycle threshold value*" OR "spread period"~5 OR "infectivity period"~5 OR "transmissability period"~5 OR "transmissability duration"~5 OR "transmission period"~5 OR "transmission duration"~5 OR "communicable period"~3 OR "contagion period" OR "communicability period"~3 OR "infectiousness period"~3 OR "contagiousness period"~3 OR "communicability period"~3 OR "shed duration"~5 OR "shedding duration"~5 OR "CT value" OR "cycle threshold values" OR "CT values" OR "cycle threshold values" OR "viral culture" OR "viral cultures" OR "negative culture" OR "viral kinetics"~5 OR "viral time"~5) |
| 2 | ("infective length"~5 OR "contagious length"~5 OR "infectious length"~5 OR "spread length"~5 OR "infectivity length"~5 OR "infective time"~5 OR "contagious time"~5 OR "infectious time"~5 OR "spread time"~5 OR "infectivity time"~5 OR "transmissability length"~5 OR "transmissability time"~5 OR "transmission length"~5 OR "transmission time"~5 OR "shed length"~5 OR "shedding length"~5 OR "shed time"~5 OR "shedding time"~5 OR "communicability length"~5 OR "communicability time"~5) |
| 3 | ti:("infective duration"~5 OR "contagious duration"~5 OR "infectious duration"~5 OR "CT value*" OR "cycle threshold value*" OR "spread duration"~5 OR "infectivity duration"~5 OR "infective period"~5 OR "contagious period"~5 OR "infectious period"~5 OR "spread period"~5 OR "infectivity period"~5 OR "transmissability period"~5 OR "transmissability duration"~5 OR "transmission period"~5 OR "transmission duration"~5 OR "shed duration"~5 OR "shedding duration"~5 OR "CT value" OR "cycle threshold values" OR "CT values" OR "cycle threshold values" OR "infective length"~5 OR "contagious length"~5 OR "infectious length"~5 OR "spread length"~5 OR "infectivity length"~5 OR "infective time"~5 OR "contagious time"~5 OR "infectious time"~5 OR "spread time"~5 OR "infectivity time"~5 OR "transmissability length"~5 OR "transmissability time"~5 OR "transmission length"~5 OR "transmission time"~5 OR "shed length"~5 OR "shedding length"~5 OR "shed time"~5 OR "shedding time"~5 OR "communicability length"~5 OR "communicability time"~5) |
| 4 | #1 OR #2 OR #3 |
| 5 | #4 AND entry_date:([20220101 TO 20220728]) |
| 6 | ((*isolat* OR quarantin*) AND ("antigen test"~3 OR "antigen tests"~3 OR "antigen testing"~3 OR "rapid test"~5 OR "rapid tests"~5 OR "rapid testing"~3 OR "lateral test"~5 OR "lateral tests"~5 OR "lateral testing"~5 OR "lateral flow antigen"~5) ) AND entry_date:([20211123 TO 20220728]) |
| 7 | #5 OR #6 |

### Appendix 2: Methods of the complex microsimulation model

Quilty and colleagues developed a stochastic, individual-based model of different isolation and testing strategies for reducing onward transmission from COVID-19 patients.^1^ We adopted this model and revised it to suit our study purposes. We used data specific to the Omicron variant when available to parameterize the model. Key model parameters are presented in the table below. Using a baseline Ct level of 40, incubation period of 3.42 days,^2^ a peak Ct value of 22.3 for symptomatic individuals,^3^ viral shedding time for symptomatic infections of 19.7 days (95%CI 17.2 to 22.7) and asymptomatic infections of 10.9 days (95% CI 8.3 to 14.3 days),^4^ and day 5, day 6, and day 10 viral culture positivity or rapid antigen test positivity data from our rapid systematic review, we simulated a more accurate viral load trajectory of Ct values over the course of infection for each individual. We assumed that if the Ct value is less than 30, the individual is infectious.^5^

We then applied different isolation strategies and simulated a sample of 1000 individuals. We assumed the isolation adherence is 100% and in the daily rapid testing strategy, the test adherence is 100%. We sampled the proportion of asymptomatic index cases from a beta distribution with a median of 31% (95%CI 24% to 38%).^6^ To estimate secondary cases, we used the basic reproduction number of 2.66 (95% CI 2.41 to 2.94) from a meta-analysis.^7^ We used the 95% CI of the basic reproduction number to calculate 95% uncertainty intervals (UIs). To estimate onward transmission leading to hospitalization and death, we sampled hospitalization rate from a beta distribution with a median of 5.54% (95%CI 3.30% to 6.00%) and sampled mortality from a beta distribution with a median of 1.33% (95%CI 0% to 2.00%).^8^

For the isolation of five days and ten days, we estimated hospitalization and death for secondary cases using both the rapid antigen test data and viral culture data for the model. For the removal of isolation based on a negative antigen test, to estimate hospitalization and death for secondary cases, we compared the difference between the trajectory of Ct values based on rapid antigen test data and Ct values simulated by viral culture data.

**Table. Key parameters used for the complex microsimulation model**

| **Parameter** | **Value** | **Source** |
| --- | --- | --- |
| Viral culture positivity on day 5 | beta distribution, 63.64%, 95%CI 32.69% to 90% | our rapid review |
| Viral culture positivity on day 6 | beta distribution, 35.28%, 95%CI 5.04% to 73.51% | our rapid review |
| Viral culture positivity on day 10 | beta distribution, 0%, 95%CI 0% to 15.07% | our rapid review |
| Rapid antigen test positivity on day 5 | beta distribution, 48.26%, 95%CI 34.15% to 62.5% | our rapid review |
| Rapid antigen test positivity on day 6 | beta distribution, 47.45%, 95%CI 28.2% to 67.09% | our rapid review |
| Rapid antigen test positivity on day 10 | beta distribution, 21.46%, 95%CI 0% to 64.14% | our rapid review |
| Incubation period | log-normal (log-mean 1.12, log-SD 0.41), mean 3.42 days, 95% CI 2.88 to 3.96 days | Wu 2022^2^ |
| Peak Ct value for symptomatic individuals | normally distributed, mean 22.3, SD 4.2 | Kissler 2021^3^ |
| Ct value threshold for an individual that is infectious | Ct value < 30 | Singanayagam 2020^5^ |
| Isolation adherence | 100% | assumed |
| Rapid antigen test adherence | 100% | assumed |
| Duration of viral shedding | symptomatic infections: mean 19.7 days, 95%CI 17.2 to 22.7 days; asymptomatic infections: mean 10.9 days, 95% CI 8.3 to 14.3 days | Yan 2021^4^ |
| Asymptomatic fraction of index cases | beta distribution, 31%, 95% CI 24% to 38% | Buitrago-Garcia 2020^6^ |
| Basic reproduction number | 2.66, 95% CI 2.41 to 2.94 | Dhungel 2022^7^ |
| Hospitalization rate | beta distribution, 5.44%, 95%CI 3.30% to 6.00% | Pitre 2022^8^ |
| Mortality | beta distribution, 1.33%, 95%CI 0 to 2.00% | Pitre 2022^8^ |

### Appendix 3. Risk of bias for clinical studies

| **Study** | **Study** **participation** | **Study attrition** | **Prognostic factor measurement** | **Outcome measurement** | **Statistical analysis and reporting** | **Overall** |
| --- | --- | --- | --- | --- | --- | --- |
| Alshukairi 2022 | Low | Low | Moderate | Low | Low | Low |
| Bouton 2022 | Low | Low | Moderate | Low | Low | Low |
| Cosimi 2022 | Low | Low | Low | Low | Moderate | Low |
| Côté 2022 | Low | Low | Moderate | Low | Moderate | Moderate |
| Earnest 2022 | Low | Low | Low | Low | Low | Low |
| Jang 2022 | Low | Low | Moderate | Low | Moderate | Moderate |
| Landon 2022 | Low | Low | Moderate | Low | Moderate | Moderate |
| Lefferts 2022 | Low | Low | Low | Low | Moderate | Low |
| Mack 2022 | Low | Low | Moderate | Low | Moderate | Moderate |
| Nelson 2022 | Low | Low | Low | Low | Low | Low |
| Sikka 2022 | Low | Low | Moderate | Low | Low | Low |
| Stingone 2022 | Low | Low | Low | Low | Low | Low |

### Appendix 4. Pooled percentage of rapid antigen test positivity for overall patients

#### 4.1. Pooled percentage of rapid antigen test positivity from day 5 to day 8 for overall patients

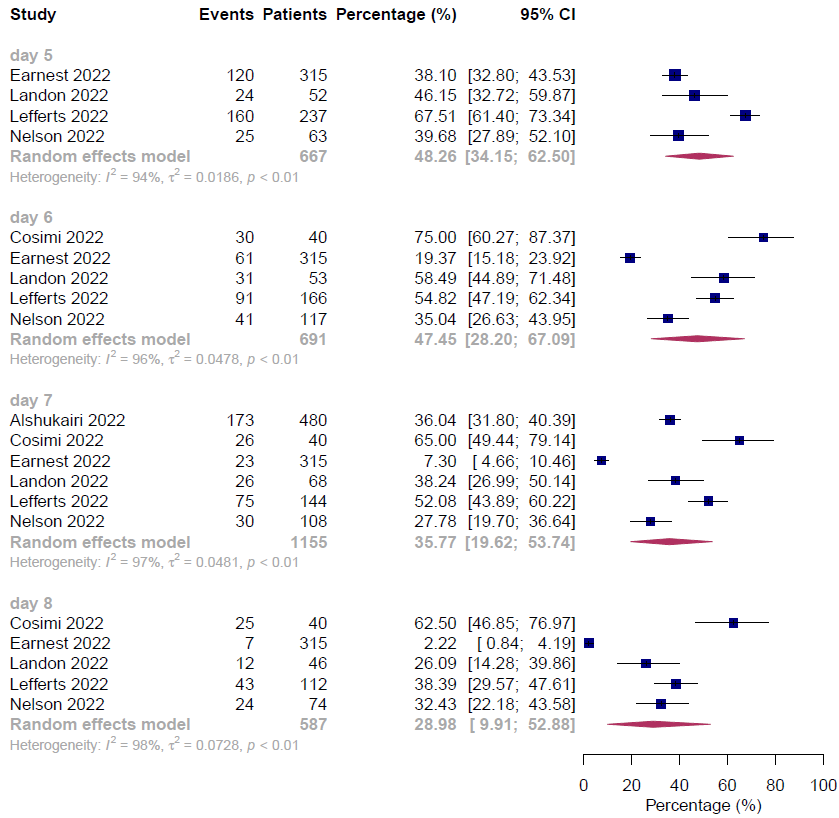

#### 4.2. Pooled percentage of rapid antigen test positivity from day 9 to day 14 for overall patients

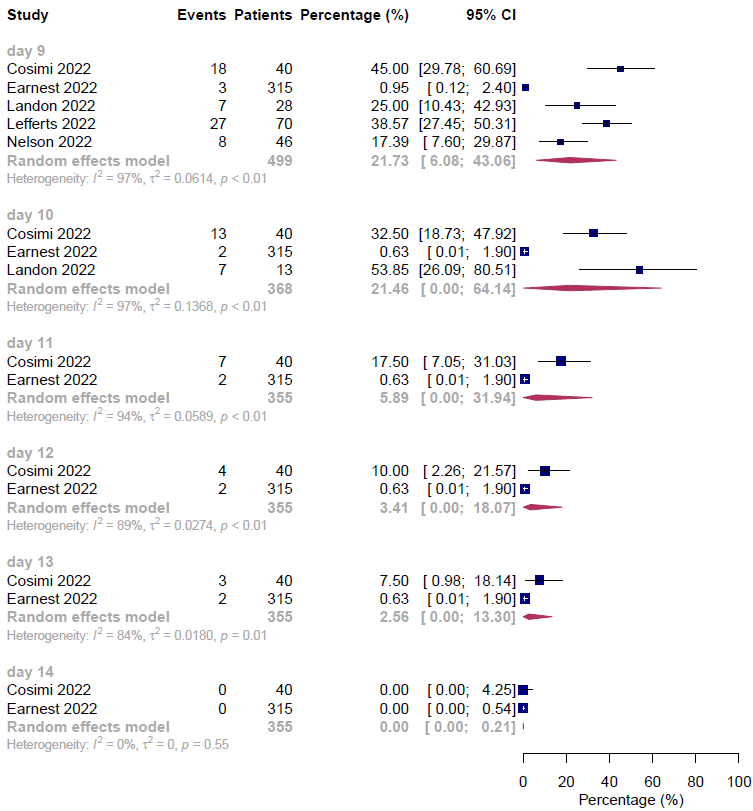

### Appendix 5. GRADE assessment for rapid antigen test positivity and viral culture positivity at days 5, 6 and 10

| **Outcomes** | **№ of studies** | **Effect** | | | **Certainty of the evidence** |
| --- | --- | --- | --- | --- | --- |
|  |  | **№ of events** | **№ of individuals** | **Percentage**  **(95% CI)** |  |
| Day 5 rapid antigen test positivity | 4 | 329 | 667 | pooled percentage  48.26 per 100 (34.15 to 62.5) | ⨁⨁⨁◯  Moderate† |
| Day 6 rapid antigen test positivity | 5 | 254 | 691 | pooled percentage  47.45 per 100 (28.20 to 67.09) | ⨁⨁⨁◯  Moderate§ |
| Day 10 rapid antigen test positivity | 3 | 22 | 368 | pooled percentage  21.46 per 100 (0 to 64.14) | ⨁⨁⨁◯  Moderate§ |
| Day 5 viral culture positivity | 1 | 7 | 11 | pooled percentage  63.64 per 100 (32.69 to 90.00) | ⨁⨁◯◯  Low* |
| Day 6 viral culture positivity | 3 | 24 | 120 | pooled percentage  35.28 per 100 (5.04 to 73.51) | ⨁⨁◯◯  Low†‡ |
| Day 10 viral culture positivity | 1 | 0 | 11 | pooled percentage  0 per 100 (0 to 15.07) | ⨁⨁◯◯  Low* |

†Rated down one level for inconsistency due to the high heterogeneity.

§Rated down one level for a combination of inconsistency and imprecision.

*Rated down two levels for imprecision due to the wide 95% confidence interval and small sample size.

‡Rated down one level for imprecision due to the wide 95% confidence interval.

### Appendix 6. Within-study subgroup analysis of day 7, day 8 and day 9 rapid antigen test positivity by symptom status

#### 6.1. Within-study subgroup analysis of day 7 rapid antigen test positivity by symptom status

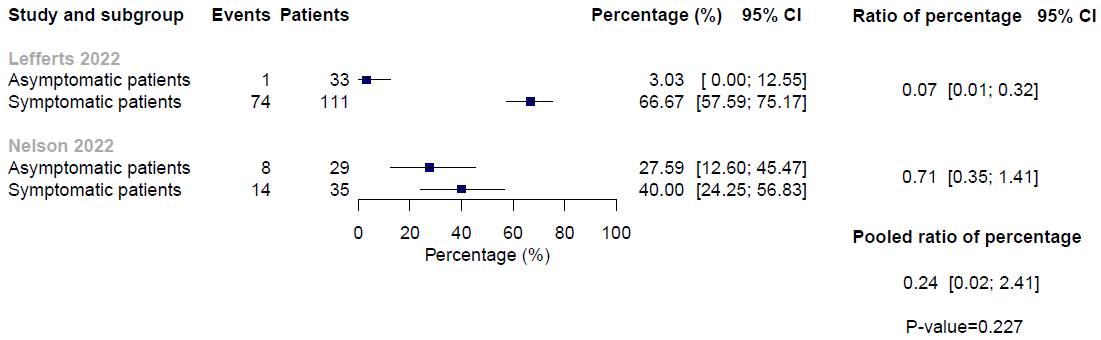

#### 6.2. Within-study subgroup analysis of day 8 rapid antigen test positivity by symptom status

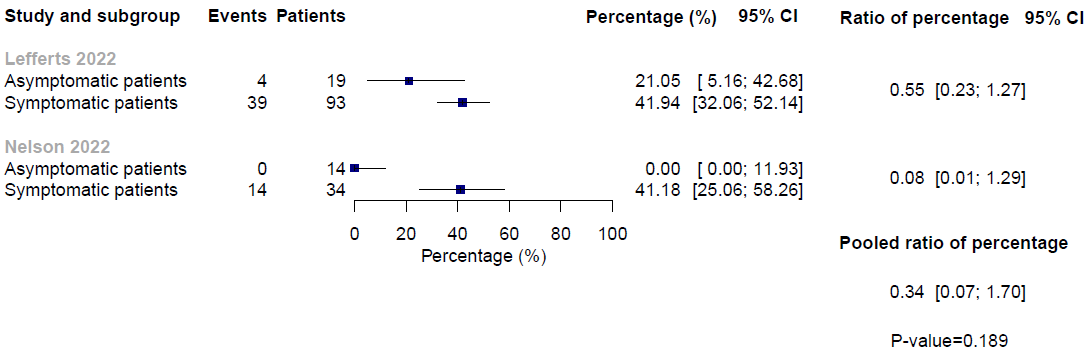

#### 6.3. Within-study subgroup analysis of day 9 rapid antigen test positivity by symptom status

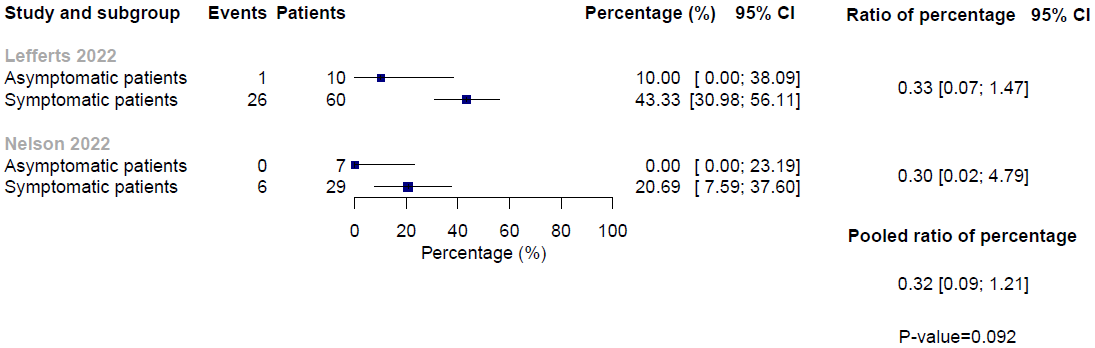

### Appendix 7. Pooled percentage of viral culture positivity from day 5 to day 14

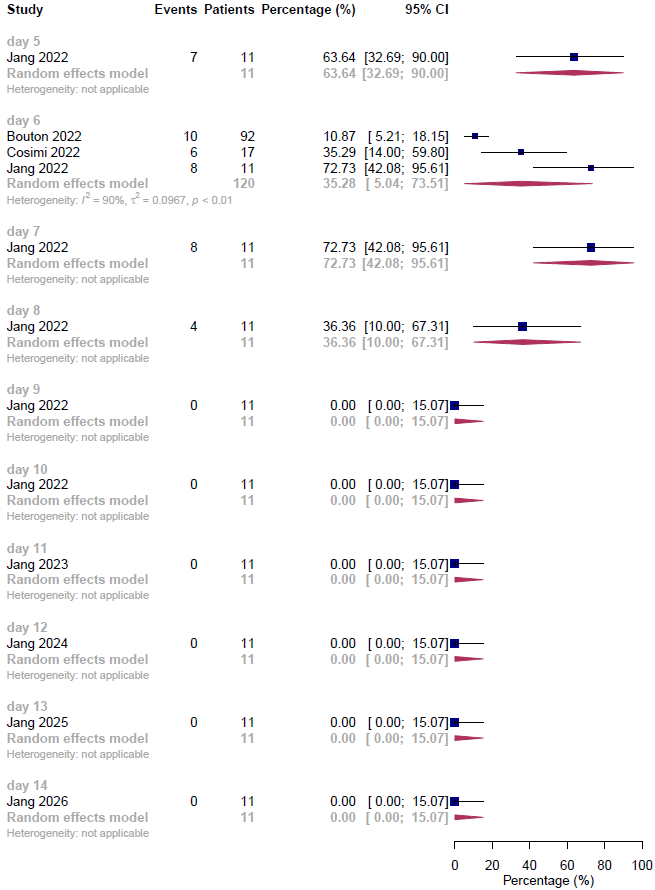

### Appendix 8. Subgroup analysis of day 6 viral culture positivity

#### 8.1. Subgroup analysis of day 6 viral culture positivity by symptom status

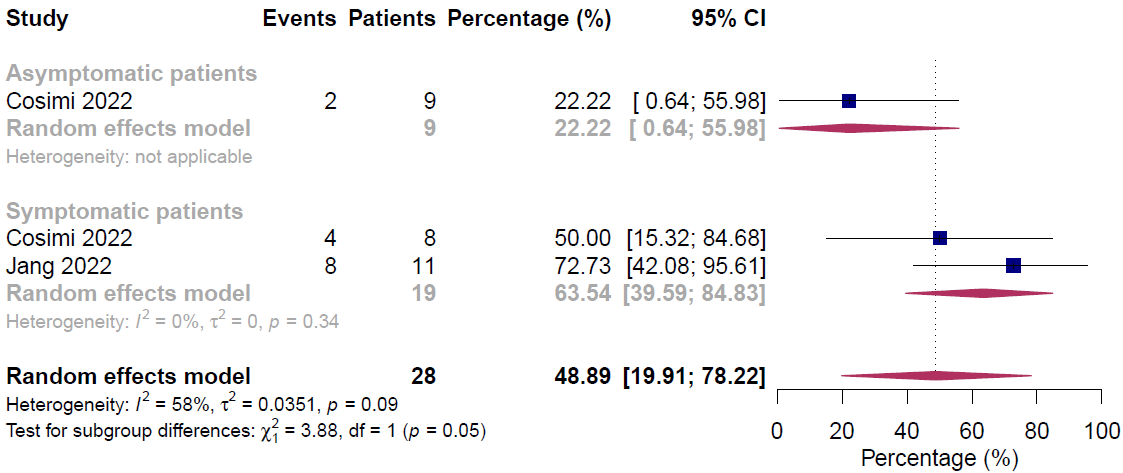

#### 8.2. Subgroup analysis of day 6 viral culture positivity by vaccination status

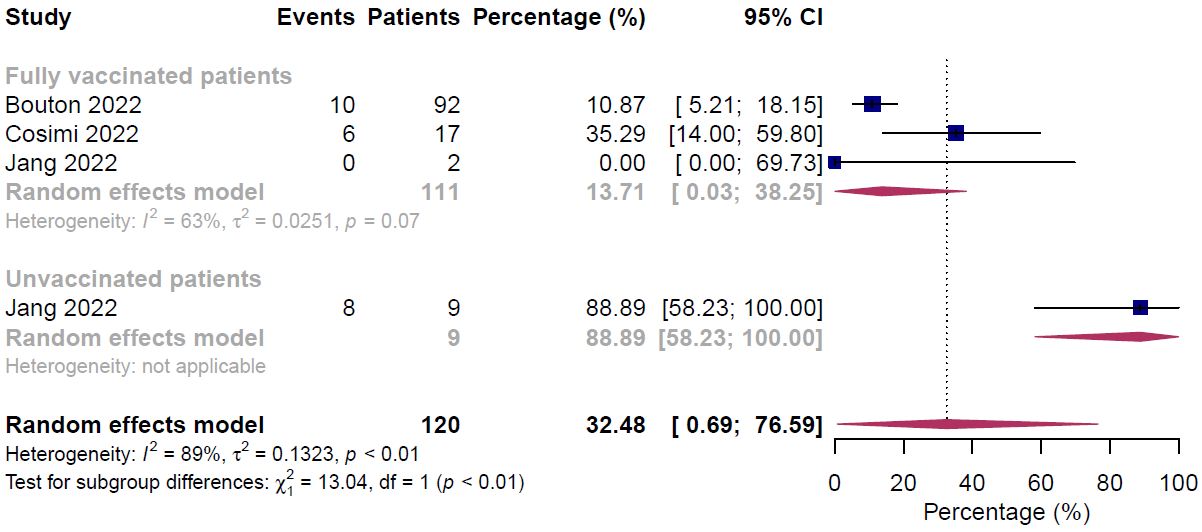

### Appendix 9. Credibility assessment of subgroup analysis for day 6 viral culture positivity

#### 9.1. Credibility assessment of subgroup analysis for day 6 viral culture positivity by symptom status

| **Credibility assessment** | | | |
| --- | --- | --- | --- |
| **1: Is the analysis of effect modification based on comparison within rather than between trials?** | | | |
| [ ] Completely between | [ **X** ] Mostly between or unclear | [ ] Mostly within | [ ] Completely within |
| *Subgroup analysis or meta-regression comparing overall effects of each individual trial. This is typical for aggregate data meta-analysis.* | *Subgroup analysis or meta-regression with most information coming from overall effects, but some trials providing within-trial subgroup information* | *Most trials providing within-trial subgroup information; or individual participant data analysis that combines within and between trial information* | *All trials providing within-trial subgroup information or individual participant data; and the analysis separates within from between trial information, e.g., meta-analysis of interactions* |
| Comment: 1 study provided within study information. | | | |
| **2: For within-trial comparisons, is the effect modification similar from trial to trial?** [ **X** ] Not applicable: no or one within-RCT comparison | | | |
| [ ] Definitely not similar | [ ] Probably not similar or unclear | [ ] Mostly similar | [ ] Definitely similar |
| *Effect modification reported for two or more trials and clearly different directions* | *Effect modification not reported for individual trials or too imprecise to tell* | *Effect modification reported for two or more trials, mostly similar in direction, but considerable differences in magnitude* | *Effect modification reported for two or more trials, similar in direction, only some differences in magnitude* |
| Comment: 1 study provided within study information. | | | |
| **3: For between-trial comparisons, is the number of trials large?** [ ] Not applicable: no between RCT comparison | | | |
| [ **X** ] Very small | [ ] Rather small or unclear | [ ] Rather large | [ ] Large |
| *1 or 2 or in smallest subgroup; 5 or less in continuous meta-regression* | *3-4 in smallest subgroup; 6-10 in continuous meta-regression* | *5-9 in smallest subgroup; 11 to 15 in continuous meta-regression* | *10 or more in smallest subgroup; more than 15 in continuous meta-regression* |
| Comment: 1 study in smallest subgroup. | | | |
| **4: Was the direction of effect modification correctly hypothesized a priori?** | | | |
| [ ] Definitely no | [ ] Probably no or unclear | [ ] Probably yes | [ **X** ] Definitely yes |
| *Clearly post-hoc or results inconsistent with hypothesized direction or biologically very implausible* | *Vague hypothesis or hypothesized direction unclear* | *No prior protocol available but unequivocal statement of a priori hypothesis with correct direction of effect modification* | *Prior protocol available and includes correct specification of direction of effect modification, e.g., based on a biologic rationale* |
| Comment: Positivity rate is lower in asymptomatic patients. | | | |
| **5: Does a test for interaction suggest that chance is an unlikely explanation of the apparent effect modification?** (consider irrespective of number of effect modifiers) | | | |
| [ ] Chance a very likely explanation | [ **X** ] Chance a likely explanation or unclear | [ ] Chance may not explain | [ ] Chance an unlikely explanation |
| *Interaction or meta-regression p-value >0.05* | *Interaction or meta-regression p-value ≤0.05 and >0.01, or no test of interaction reported and not computable* | *Interaction or meta-regression p-value ≤0.01 and >0.005* | *Interaction or meta-regression p-value ≤0.005* |
| Comment: P=0.05 | | | |
| **6: Did the authors test only a small number of effect modifiers or consider the number in their statistical analysis?** | | | |
| [ ] Definitely no | [ ] Probably no or unclear | [ ] Probably yes | [ **X** ] Definitely yes |
| *Explicitly exploratory analysis or large number of effect modifiers tested (e.g., greater than 10) and multiplicity not considered in analysis* | *No mention of number or 4-10 effect modifiers tested and number not considered in analysis* | *No protocol available but unequivocal statement of 3 or fewer effect modifiers tested* | *Protocol available and 3 or fewer effect modifiers tested or number considered in analysis* |
| Comment: Tested 2 effect modifiers. | | | |
| **7: Did the authors use a random effects model?** [ ] Not applicable | | | |
| [ ] Definitely no | [ ] Probably no or unclear | [ ] Probably yes | [ **X** ] Definitely yes |
| *Fixed (or common) effect or fixed effects model explicitly stated* | *Probably fixed effect(s) model* | *Probably random (or mixed) effects* | *Random (or mixed) effects explicitly stated* |
| Comment: Random effects model. | | | |
| **8: If the effect modifier is a continuous variable, were arbitrary cut points avoided?** [ **X** ] not applicable: not continuous | | | |
| [ ] Definitely no | [ ] Probably no or unclear | [ ] Probably yes | [ ] Definitely yes |
| *Analysis based on exploratory cut point(s), e.g., picking cut point associated with highest interaction p-value* | *Analysis based on cut point(s) of unclear origin* | *Analysis based on pre-specified cut point(s), e.g., suggested by prior RCT* | *Analysis based on the full continuum, e.g., assuming a linear or logarithmic relationship* |
| Comment: | | | |
| **9 Optional: Are there any additional considerations that may increase or decrease credibility?** (manual section 3.9) [ **X** ] not applicable | | | |
|  | [ ] Yes, probably decrease  Biologically implausible  Expect similar severe critical  Opposite effects unlikely | [ ] Yes, probably increase | |
| Comment: The cut point for categorization appears to be data driven The number of events driving the p-value is extremely small Biology seems very dubious **10: How would you rate the overall credibility of the proposed effect modification?** The overall rating should be driven by the items that decrease credibility. The following provides a sensible strategy: - All responses definitely or probably decrease credibility or unclear 🡪 very low - Two or more responses definitely decrease credibility 🡪 maximum usually low even if all other responses satisfy credibility criteria - One response definitely decreases credibility 🡪 maximum usually moderate even if all other responses satisfy credibility criteria - Two responses probably decrease credibility 🡪 maximum usually moderate even if all other responses satisfy credibility criteria - No response options definitely or probably decrease credibility 🡪 high very likely Place a mark on the continuous line (or type “x” in editable version) \| \| \| \| \|; \| **X** \| \| \| \|; \| **Very low credibility** \| **Low credibility** \| **Moderate credibility** \| **High credibility** \|; \| Very likely no effect modification Use overall effect for each subgroup \| Likely no effect modification Use overall effect for each subgroup but note remaining uncertainty \| Likely effect modification Use separate effects for each subgroup but note remaining uncertainty \| Very likely effect modification Use separate effects for each subgroup \|; Comment: Between-trial comparison, the number of studies is small in smallest subgroup, and chance remains a likely explanation for the finding. \| \| \| \| \| | | | |

#### 9.2. Credibility assessment of subgroup analysis for day 6 viral culture positivity by vaccination status

| **Credibility assessment** | | | |
| --- | --- | --- | --- |
| **1: Is the analysis of effect modification based on comparison within rather than between trials?** | | | |
| [ ] Completely between | [ **X** ] Mostly between or unclear | [ ] Mostly within | [ ] Completely within |
| *Subgroup analysis or meta-regression comparing overall effects of each individual trial. This is typical for aggregate data meta-analysis.* | *Subgroup analysis or meta-regression with most information coming from overall effects, but some trials providing within-trial subgroup information* | *Most trials providing within-trial subgroup information; or individual participant data analysis that combines within and between trial information* | *All trials providing within-trial subgroup information or individual participant data; and the analysis separates within from between trial information, e.g., meta-analysis of interactions* |
| Comment: 1 study provided within study information. | | | |
| **2: For within-trial comparisons, is the effect modification similar from trial to trial?** [ **X** ] Not applicable: no or one within-RCT comparison | | | |
| [ ] Definitely not similar | [ ] Probably not similar or unclear | [ ] Mostly similar | [ ] Definitely similar |
| *Effect modification reported for two or more trials and clearly different directions* | *Effect modification not reported for individual trials or too imprecise to tell* | *Effect modification reported for two or more trials, mostly similar in direction, but considerable differences in magnitude* | *Effect modification reported for two or more trials, similar in direction, only some differences in magnitude* |
| Comment: 1 study provided within study information. | | | |
| **3: For between-trial comparisons, is the number of trials large?** [ ] Not applicable: no between RCT comparison | | | |
| [ **X** ] Very small | [ ] Rather small or unclear | [ ] Rather large | [ ] Large |
| *1 or 2 or in smallest subgroup; 5 or less in continuous meta-regression* | *3-4 in smallest subgroup; 6-10 in continuous meta-regression* | *5-9 in smallest subgroup; 11 to 15 in continuous meta-regression* | *10 or more in smallest subgroup; more than 15 in continuous meta-regression* |
| Comment: 1 study in smallest subgroup. | | | |
| **4: Was the direction of effect modification correctly hypothesized a priori?** | | | |
| [ ] Definitely no | [ ] Probably no or unclear | [ ] Probably yes | [ **X** ] Definitely yes |
| *Clearly post-hoc or results inconsistent with hypothesized direction or biologically very implausible* | *Vague hypothesis or hypothesized direction unclear* | *No prior protocol available but unequivocal statement of a priori hypothesis with correct direction of effect modification* | *Prior protocol available and includes correct specification of direction of effect modification, e.g., based on a biologic rationale* |
| Comment: Positivity rate is lower in fully vaccinated patients. | | | |
| **5: Does a test for interaction suggest that chance is an unlikely explanation of the apparent effect modification?** (consider irrespective of number of effect modifiers) | | | |
| [ ] Chance a very likely explanation | [ ] Chance a likely explanation or unclear | [ **X** ] Chance may not explain | [ ] Chance an unlikely explanation |
| *Interaction or meta-regression p-value >0.05* | *Interaction or meta-regression p-value ≤0.05 and >0.01, or no test of interaction reported and not computable* | *Interaction or meta-regression p-value ≤0.01 and >0.005* | *Interaction or meta-regression p-value ≤0.005* |
| Comment: P<0.01 | | | |
| **6: Did the authors test only a small number of effect modifiers or consider the number in their statistical analysis?** | | | |
| [ ] Definitely no | [ ] Probably no or unclear | [ ] Probably yes | [ **X** ] Definitely yes |
| *Explicitly exploratory analysis or large number of effect modifiers tested (e.g., greater than 10) and multiplicity not considered in analysis* | *No mention of number or 4-10 effect modifiers tested and number not considered in analysis* | *No protocol available but unequivocal statement of 3 or fewer effect modifiers tested* | *Protocol available and 3 or fewer effect modifiers tested or number considered in analysis* |
| Comment: Tested 2 effect modifiers. | | | |
| **7: Did the authors use a random effects model?** [ ] Not applicable | | | |
| [ ] Definitely no | [ ] Probably no or unclear | [ ] Probably yes | [ **X** ] Definitely yes |
| *Fixed (or common) effect or fixed effects model explicitly stated* | *Probably fixed effect(s) model* | *Probably random (or mixed) effects* | *Random (or mixed) effects explicitly stated* |
| Comment: Random effects model. | | | |
| **8: If the effect modifier is a continuous variable, were arbitrary cut points avoided?** [ **X** ] not applicable: not continuous | | | |
| [ ] Definitely no | [ ] Probably no or unclear | [ ] Probably yes | [ ] Definitely yes |
| *Analysis based on exploratory cut point(s), e.g., picking cut point associated with highest interaction p-value* | *Analysis based on cut point(s) of unclear origin* | *Analysis based on pre-specified cut point(s), e.g., suggested by prior RCT* | *Analysis based on the full continuum, e.g., assuming a linear or logarithmic relationship* |
| Comment: | | | |
| **9 Optional: Are there any additional considerations that may increase or decrease credibility?** (manual section 3.9) [ **X** ] not applicable | | | |
|  | [ ] Yes, probably decrease  Biologically implausible  Expect similar severe critical  Opposite effects unlikely | [ ] Yes, probably increase | |
| Comment: The cut point for categorization appears to be data driven The number of events driving the p-value is extremely small Biology seems very dubious **10: How would you rate the overall credibility of the proposed effect modification?** The overall rating should be driven by the items that decrease credibility. The following provides a sensible strategy: - All responses definitely or probably decrease credibility or unclear 🡪 very low - Two or more responses definitely decrease credibility 🡪 maximum usually low even if all other responses satisfy credibility criteria - One response definitely decreases credibility 🡪 maximum usually moderate even if all other responses satisfy credibility criteria - Two responses probably decrease credibility 🡪 maximum usually moderate even if all other responses satisfy credibility criteria - No response options definitely or probably decrease credibility 🡪 high very likely Place a mark on the continuous line (or type “x” in editable version) \| \| \| \| \|; \| **X** \| \| \| \|; \| **Very low credibility** \| **Low credibility** \| **Moderate credibility** \| **High credibility** \|; \| Very likely no effect modification Use overall effect for each subgroup \| Likely no effect modification Use overall effect for each subgroup but note remaining uncertainty \| Likely effect modification Use separate effects for each subgroup but note remaining uncertainty \| Very likely effect modification Use separate effects for each subgroup \|; Comment: Between-trial comparison, the number of studies is small in smallest subgroup. \| \| \| \| \| | | | |

### Appendix 10. Additional results of rapid systematic review

In one study^1^ with 173 fully vaccinated patients, 45.66% of patients had at least one negative or Ct ≥ 35 RT-PCR test result on or before day 6 and the median time from diagnosis to first negative result was 7 days (IQR 5 to 9 days). In one study^2^ with 37 patients, the percentage of PCR-positive with Ct value < 30 from day 5 to day 10 varied from 51.35% to 8.11% and the average time to PCR clearance was 7.94 days. One study^3^ with 196 SARS-CoV-2 positive homeless people reported that 26.53% of patients had a positive PCR test after 21 days of isolation and the median duration of positivity was 21 days (IQR 14 to 26).

### Appendix 11. GRADE summary of findings for five-day isolation versus ten-day isolation for outcomes estimated using positive viral culture data from relatively simple model

| **Outcome** | **Absolute effect estimates** | | **Certainty of the evidence** | **Plain language summary** |
| --- | --- | --- | --- | --- |
|  | Isolation for 5 days | Isolation for 10 days |  |  |
| **We assume all patients with** **positive** **viral culture are infectious** | | | | |
| Onward transmission leading to hospitalization (28 days) | **26**  per 1000 | **0**  per 1000 | **Very low**  Due to certainty of parameters (low) in the model and indirectness | Whether isolation of 5 days compared with 10 days would increase hospitalization for secondary cases is very uncertain. |
|  | Difference: **26 more per 1000**  (95% UI 20 more to 33 more) | |  |  |
| Onward transmission leading to death (90 days) | **6**  per 1000 | **0**  per 1000 | **Very low**  Due to certainty of parameters (low) in the model and indirectness | Whether isolation of 5 days compared with 10 days would increase mortality for secondary cases is very uncertain. |
|  | Difference: **6 more per 1000**  (95% UI 5 more to 8 more) | |  |  |

UI, uncertainty interval.

### Appendix 12. GRADE summary of findings for estimates from complex microsimulation model for overall patients

#### 12.1. GRADE summary of findings for five-day isolation versus ten-day isolation for estimates from complex microsimulation model

| **Outcome** | **Absolute effect estimates** | | **Certainty of the evidence** | **Plain language summary** |
| --- | --- | --- | --- | --- |
|  | Isolation for 5 days | Isolation for 10 days |  |  |
| **Use rapid antigen test data for the model** | | | | |
| Onward transmission leading to hospitalization (28 days) | **14**  per 1000 | **1**  per 1000 | **Very low**  Due to certainty of parameters (moderate) in the model and indirectness | Whether isolation of 5 days compared with 10 days would increase hospitalization for secondary cases is very uncertain. |
|  | Difference: **13 more per 1000**  (95% UI 12 more to 16 more) | |  |  |
| Onward transmission leading to death (90 days) | **7**  per 1000 | **0**  per 1000 | **Very low**  Due to certainty of parameters (moderate) in the model and indirectness | Whether isolation of 5 days compared with 10 days would increase mortality for secondary cases is very uncertain. |
|  | Difference: **7 more per 1000**  (95% UI 6 more to 7 more) | |  |  |
| **Use viral culture data for the model** | | | | |
| Onward transmission leading to hospitalization (28 days) | **16**  per 1000 | **0**  per 1000 | **Very low**  Due to certainty of parameters (low) in the model and indirectness | Whether isolation of 5 days compared with 10 days would increase hospitalization for secondary cases is very uncertain. |
|  | Difference: **16 more per 1000**  (95% UI 13 more to 17 more) | |  |  |
| Onward transmission leading to death (90 days) | **7**  per 1000 | **0**  per 1000 | **Very low**  Due to certainty of parameters (low) in the model and indirectness | Whether isolation of 5 days compared with 10 days would increase mortality for secondary cases is very uncertain. |
|  | Difference: **7 more per 1000**  (95% UI 5 more to 7 more) | |  |  |

UI, uncertainty interval.

#### 12.2. GRADE summary of findings for removal of isolation based on a negative antigen test versus ten-day isolation for estimates from complex microsimulation model

| **Outcome** | **Absolute effect estimates** | | **Certainty of the evidence** | **Plain language summary** |
| --- | --- | --- | --- | --- |
|  | Removal of isolation based on a negative antigen test | Isolation for 10 days |  |  |
| Onward transmission leading to hospitalization (28 days) | **14**  per 1000 | **0**  per 1000 | **Very low**  Due to certainty of parameters (low) in the model and indirectness | Whether removing isolation based on a negative antigen test compared with isolation of 10 days would increase hospitalization for secondary cases is very uncertain. |
|  | Difference: 14 **more per 1000**  (95% UI 12 more to 14 more) | |  |  |
| Onward transmission leading to death (90 days) | **5**  per 1000 | **0**  per 1000 | **Very low**  Due to certainty of parameters (low) in the model and indirectness | Whether removing isolation based on a negative antigen test compared with isolation of 10 days would increase mortality for secondary cases is very uncertain. |
|  | Difference: **5 more per 1000**  (95% UI 5 more to 6 more) | |  |  |

UI, uncertainty interval.
